# Association of Cardiovascular Manifestations with In-hospital Outcomes in Patients with COVID-19: A Hospital Staff Data

**DOI:** 10.1101/2020.02.29.20029348

**Authors:** Ru Liu, Xiaoyan Ming, Ou Xu, Jianli Zhou, Hui Peng, Ning Xiang, Jiaming Zhang, Hong Zhu

## Abstract

**Background:** The outbreaks of coronavirus disease 2019 (COVID-19) caused by the novel severe acute respiratory syndrome coronavirus 2 (SARS-CoV-2) remain a huge threat to the public health worldwide. Clinical data is limited up to now regarding the risk factors in favor of severe conversion of non-severe case with COVID-19.

**Aims:** This study analyzed a hospital staff data to figure out general clinical features of COVID-19 in terms of the association of cardiovascular manifestations (CVMs) with in-hospital outcomes of COVID-19 cases.

**Methods:** Retrospective, single-center case series of 41 consecutive hospitalized health staff with confirmed COVID-19 were collected at the Central Hospital of Wuhan in Wuhan, China, from January 15 to January 24, 2020. Epidemiological, demographic, clinical, laboratory, radiological, treatment data and in-hospital adverse events were collected and analyzed. Final date of follow-up was March 3, 2020. A comparative study was applied between cases with CVMs and those without CVMs.

**Results:** Of all, clinicians and clinical nurses accounted for 80.5%, while 87.8% of all had history of patient contact. The population was presented with a mean age of 39.1 ± 9.2 and less comorbidities than community population. The three most frequent symptoms of COVID-19 cases analyzed were fever (82.9%), myalgia or fatigue (80.5%) and cough (63.4%). While, the three most frequent initial symptoms were myalgia or fatigue (80.5%), fever (73.2%) and cough (41.5%). There were 95.1% cases featured as non-severe course of disease according to the official standard in China. Patients with CVMs and those without CVMs accounted for 58.5% and 41.5%, respectively. Compared with cases without CVMs, patients with CVMs were presented with lower baseline lymphocyte count (0.99 ± 0.43 and 1.55 ± 0.61, *P*<0.001), more who had at least once positive nucleic acid detection of throat swab during admission (50.0% and 11.8%, *P*=0.011), and more received oxygen support (79.2% and 23.5%, *P*<0.001). The rate of in-hospital adverse events was significantly higher in patients with CVMs group (75.0% and 23.5%, *P*=0.001). Multivariable logistic regression model indicated that, coexisting with CVMs in COVID-19 patients was not independently associated with in-hospital adverse events.

**Conclusions:** Most of hospital staff with COVID-19 had history of patient contact, featured non-severe course of disease. Cases with CVMs suffered from more in-hospital adverse events than those without CVMs. But concomitant CVMs were not independently associated with in-hospital adverse events in COVID-19 patients.

## Introduction

Ever since December 2019, novel coronavirus pneumonia (NCP), which firstly appeared in Wuhan and then rapidly spread nationwide and abroad, has taken 2946 lives and confirmedly infected 80304 cases in China as of March 3, 2020. Outside of China, a total of 10566 cases confirmedly infected and 166 deaths have been reported in 72 countries [1-6]. The acute respiratory infectious disease caused by infection of severe acute respiratory syndrome coronavirus 2 (SARS-CoV-2) (tentatively named 2019-nCoV since official confirmation as the pathogen of the outbreak, later formally named SARS-CoV-2 by the Coronavirus Study Group of the International Committee on Taxonomy of Viruses on February 7, 2020) was officially named as coronavirus disease 2019 (COVID-19) by World Health Organization (WHO) on February 11, 2020 [7].

As clinic experiences increases, clinicians become conscious of cardiovascular comorbidities and manifestations of COVID-19. Two reports showed that patients with COVID-19 and hypertension or coronary artery disease had worse in-hospital outcomes [8, 9]. Cardiovascular manifestations (CVMs) could be the initial presentation or appear throughout the whole course of COVID-19. Liu et al analyzed 137 cases with COVID-19 and found that patients who complains palpitation as initial symptom accounted for 7.3% [10]. The first report of 41 cases with COVID-19 admitted from December 1, 2019 to January 2, 2020, revealed acute cardiac injury occurred in 12% of patients [11]. Wang et al reported in a single-center case series involving 138 patients with COVID-19, also found that cardiovascular complications of COVID-19 were not rare, among which arrhythmia and acute cardiac injury accounted for 16.7% and 7.2%, respectively. The level of hypersensitive troponin I on admission was significantly higher in those who had been admitted to intensive care unit (ICU) than those who had not [12]. Meanwhile, Wang et al hospital-associated transmission of 2019-nCoV was suspected in about 41% of patients. And clinician and clinical nurse accounted for 29% of total, shockingly [12]. The NCP Emergency Response Epidemiology Team of China reported a total of 1716 health workers have become infected and 5 have died (0.3%) as of February 11, 2020 [13]. The detailed data are still limited, regarding the clinical manifestations, epidemiological characteristics, treatment situation and outcomes in this special population. Therefore, this study analyzed a group of hospital staff diagnosed as COVID-19 to figure out this issue. Further, a comparative study was applied to explain whether concomitant CVMs had effect on the in-hospital outcomes of COVID-19 cases.

## Methods

### Ethical Statement

As a retrospective study and data analysis were performed anonymously, the requirement for informed consent was waived. The Ethics Committees of the Central Hospital of Wuhan approved this study.

### Study Population

A group of staff of the Central Hospital of Wuhan with confirmed COVID-19 admitted to the Pneumology Department of the Central Hospital of Wuhan from January 15 to January 24, 2020, were enrolled. Oral consent was obtained from patients. The Central Hospital of Wuhan located in Wuhan, Hubei Province, the endemic areas of COVID-19, is one of the major tertiary teaching hospitals affiliated to Tongji Medical College, Huazhong University of Science and Technology, and is responsible for the treatments for COVID-19 assigned by government. All patients with COVID-19 enrolled in this study were diagnosed according to WHO interim guidance, as well as the Diagnosis and Treatment Scheme of COVID-19 (the fifth trial edition) by National Health Commission and National Administration of Traditional Chinese Medicine of China [14, 15]. The clinical outcomes (ie, in-hospital adverse events, nucleic acid detection) were monitored up to March 3, 2020, the final date of follow-up.

### Data Collection and Definitions

The medical records of patients were collected into a database by the researcher of the Central Hospital of Wuhan from electronic medical records system. Epidemiological and clinical characteristics, laboratory and radiological findings, treatment and outcomes data were first-hand obtained. The data were reviewed by a trained team of physicians. Information recorded included demographic data, medical history, exposure history, comorbidities, symptoms and signs, laboratory findings, chest computed tomographic (CT) scans, nucleic acid detection of throat swab, treatment measures and in-hospital adverse events. The date of disease onset was defined as the day when the symptom was noticed. Treatment included antiviral therapy, antibiotics or anti-fungus drugs, corticosteroid therapy, intravenous human albumin or immunoglobulin, thymosin, traditional Chinese medicine, mesenchymal stem cell therapy and respiratory support. None received kidney replacement therapy, plasma exchange or artificial liver.

Respiratory failure (RF) was diagnosed according to arterial blood gas analysis. Acute respiratory distress syndrome (ARDS) was defined according to the Berlin definition [15, 16]. Acute cardiac injury was defined if the serum levels of cardiac biomarkers (mainly creatine kinase-MB and hypersensitive cardiac troponin I) were above the 99th percentile upper reference limit, with a tendency to rise or fall, or new abnormalities were shown in electrocardiography and echocardiography during the whole course of disease [12]. Acute kidney injury (AKI) was identified according to the Kidney Disease: Improving Global Outcomes definition [17]. Acute hepatic injury was recognized when elevated level of serum aminotransferase or bilirubin was detected during hospitalization, which cannot be explained by background disease. Acute myocyte injury was recognized as myalgia or fatigue along with acute hypercreatine kinasemia (peak sCK is >or=3 x normal) while no evidence on myocardial injury [18]. Shock was identified according to treatment guideline of severe sepsis/septic shock (China) [19]. Secondary infection was diagnosed by any one of: 1, elevated neutrophil percentage or procalcitonin; 2, respiratory purulent secretion, increased coughing of phlegm or moist rales; 3, radiological features of bacteria or fungus infection. In-hospital adverse events included RF or ARDS, transfer to ICU, invasive mechanic ventilation, acute cardiac injury, AKI, acute hepatic injury, acute myocyte injury, shock, secondary infection and death. Cases with CVMs were defined by any one of these throughout the course of disease: 1, complain of palpitation or chest distress; 2, elevation of creatine kinase-MB or hypersensitive cardiac troponin I (above the 99th percentile upper reference limit); 3, new abnormalities on electrocardiography including sinus tachycardia.

### Real-Time Reverse Transcription Polymerase Chain Reaction Assay for SARS-CoV-2

Throat swab samples were collected for extracting SARS-CoV-2 ribose nucleic acid (RNA) from patients with suspected or clinical diagnosis of COVID-19 (Miraclean Technology Co., Ltd, Shenzhen, China). After collection, the throat swabs were placed into a collection tube with virus preservation solution, and total RNA was extracted using the respiratory sample RNA isolation kit (Automated Nucleic Acid Extraction System, Shanghai ZJ Bio-Tech Co., Ltd, China). The suspension was used for real-time reverse transcription polymerase chain reaction (RT-PCR) assay of SARS-CoV-2 RNA. Two target genes, including open reading frame lab (ORF1ab) and nucleocapsid protein(N), were simultaneously amplified and tested during the real-time RT-PCR assay. The real time RT-PCR assay was performed using a SARS-CoV-2 nucleic acid detection kit according to the manufacturer’s protocol (Shanghai BioGerm Medical Biotechnology Co., Ltd, China). All SARS-CoV-2 Nucleic Acid Detection Kits were provided by Wuhan Centers for Disease Control and Prevention. A cycle threshold value (Ct-value) less than 37 was defined as a positive result, while a Ct-value of 40 or more was defined as negative. A Ct-value of 37 to less than 40, required retest to confirm. Specific primers and probes for detection SARS-CoV-2 and the diagnostic criteria were referred to the recommendation by the National Institute for Viral Disease Control and Prevention (China) (http://ivdc.chinacdc.cn/kyjz/202001/t20200121_211337.html).

### Statistical Analysis

Data statistics was applied using SPSS 22.0 (IBM Corp., Armonk, New York, USA). Student’s t-tests were used to compare continuous variables, which conform to a normal distribution, while Chi-square tests were applied to compare categorical variables between the two groups. Multivariate logistic regression analyses were applied to control baseline confounders. Covariates for logistic regression were those variables with significant differences between the two groups in baseline. All P values were two sided with a significance level of 0.05. Tendency of significant difference was judged when 0.05<P<0.1.

## Results

### Epidemiological Characteristics

The study population included 41 hospitalized staff with confirmed COVID-19. The mean age was 39.1 (± 9.2) years, and 17 (41.5%) were men. The clinician and clinical nurse, who had direct contact with COVID-19 patients, accounted for 46.3% and 34.1%, respectively. Cases from medical detection departments accounted for 12.2%, who also had contact with COVID-19 patients or their specimen. Only 3 (7.3%) cases were from administrative or logistics departments, who mostly had contacted with infected colleagues. Recalling epidemiological history, 97% of all could tell how they infected. Definite history of patient contact accounted for 73.2%, while suspected history of patient contact accounted for 14.6%. Only 2 cases (4.9%) told contact with infected family members, but they had also contacted with suspected COVID-19 patients, making the clue questioning. (Table 1)

**Table 1.** Demographic, epidemiological and clinical characteristics

| Variables | All<br>(n = 41) | Cases with<br>CVMs<br>(n = 24) | Cases without<br>CVMs<br>(n = 17) | P value |
| --- | --- | --- | --- | --- |
| **Demographic characteristics** |  |  |  |  |
| Male gender, % | 17 (41.5) | 11 (45.8) | 6 (35.3) | 0.5 |
| Age, years | 39.1 ± 9.2 | 40.0 ± 10.7 | 37.9 ± 6.9 | 0.498 |
| **Epidemiological characteristics, %** |  |  |  |  |
| **Department** |  |  |  | 0.578 |
| Clinician | 19 (46.3) | 13 (54.2) | 6 (35.3) |  |
| Clinical nurse | 14 (34.1) | 7 (29.2) | 7 (41.2) |  |
| From medical detection departments | 5 (12.2) | 2 (8.3) | 3 (17.6) |  |
| From administrative or logistics departments | 3 (7.3) | 2 (8.3) | 1 (5.9) |  |
| **Epidemiology** |  |  |  | 0.793 |
| Definite history of patient contact | 30 (73.2) | 18 (75.0) | 12 (70.6) |  |
| Suspected history of patient contact | 6 (14.6) | 4 (16.7) | 2 (11.8) |  |
| Community infection by family members or others | 2 (4.9) | 1 (4.2) | 1 (5.9) |  |
| Unknown | 3 (7.3) | 1 (4.2) | 2 (11.8) |  |
| **Coexisting conditions, %** |  |  |  |  |
| Current smoker | 4 (9.8) | 3 (12.5) | 1 (5.9) | 0.482 |
| Any comorbidities | 12 (29.3) | 9 (37.5) | 3 (17.6) | 0.169 |
| Hypertension | 2 (4.9) | 2 (8.3) | 0 (0.0) | 0.222 |
| DM | 2 (4.9) | 1 (4.2) | 1 (5.9) | 0.802 |
| CAD | 1 (2.4) | 1 (4.2) | 0 (0.0) | 0.394 |
| Arrhythmia | 3 (7.3) | 3 (12.5) | 0 (0.0) | 0.13 |
| COPD | 1 (2.4) | 1 (4.2) | 0 (0.0) | 0.394 |
| Chronic liver disease | 2 (4.9) | 1 (4.2) | 1 (5.9) | 0.802 |
| Malignant tumor | 2 (4.9) | 2 (8.3) | 0 (0.0) | 0.222 |
| Rheumatic disease | 3 (7.3) | 1 (4.2) | 2 (11.8) | 0.357 |
| **Signs and symptoms** |  |  |  |  |
| Fever, % | 34 (82.9) | 21 (87.5) | 13 (76.5) | 0.355 |
| Fever as initial symptom, % | 30 (73.2) | 19 (79.2) | 11 (64.7) | 0.303 |
| Highest temperature, °C | 38.3 ± 0.7 | 38.3 ± 0.8 | 38.3 ± 0.4 | 0.763 |
| Myalgia/Fatigue, % | 33 (80.5) | 21 (87.5) | 12 (70.6) | 0.178 |
| Myalgia/Fatigue as initial symptom, % | 33 (80.5) | 21 (87.5) | 12 (70.6) | 0.178 |
| Cough, % | 26 (63.4) | 18 (75.0) | 8 (47.1) | 0.067 |
| Cough as initial symptom, % | 17 (41.5) | 11 (45.8) | 6 (35.3) | 0.5 |
| Sputum production, % | 16 (39.0) | 13 (54.2) | 3 (17.6) | 0.018 |
| Rhinobyon/running nose, % | 4 (9.8) | 4 (16.7) | 0 (0.0) | 0.076 |
| Chilly/shivering, % | 12 (29.3) | 6 (25.0) | 6 (35.3) | 0.475 |
| Headache, % | 2 (4.9) | 2 (8.3) | 0 (0.0) | 0.222 |
| Dizziness, % | 5 (12.2) | 2 (8.3) | 3 (17.6) | 0.369 |
| Hemoptysis/bloody sputum, % | 1 (2.4) | 1 (4.2) | 0 (0.0) | 0.394 |
| Diarrhea, % | 8 (19.5) | 7 (29.2) | 1 (5.9) | 0.064 |
| Somnipathy, % | 5 (12.2) | 5 (20.8) | 0 (0.0) | 0.045 |
| Wheeze, % | 14 (34.1) | 10 (41.7) | 4 (23.5) | 0.228 |
| Dyspnea, % | 12 (29.3) | 8 (33.3) | 4 (23.5) | 0.497 |
| Conjunctivitis, % | 1 (2.4) | 1 (4.2) | 0 (0.0) | 0.394 |
| Baseline RR, times/min | 19.3 ± 3.1 | 19.7 ± 3.1 | 18.8 ± 3.0 | 0.34 |
| Baseline HR, beat/min | 91.5 ± 14.6 | 96.3 ± 16.0 | 84.8 ± 8.9 | 0.01 |
| Baseline SBP, mmHg | 121.2 ± 10.5 | 123.8 ± 10.3 | 117.5 ± 9.9 | 0.055 |

CAD, coronary artery disease; COPD, chronic obstructive pulmonary disease; CVM, cardiovascular manifestation; DM, diabetes mellitus; HR, heart rate; RR, respiratory rate; SBP, systolic blood pressure
Data are expressed as mean ± standard deviation; or counts (percentage).

### Clinical Presentation, Laboratory and Radiological Data

As a young and middle-aged population, mostly of them were generally healthy, presented as less comorbidities (29.3%) than general community population previously reported [11, 12]. The comorbidities with high incidence in the elderly like diabetes mellitus (DM), hypertension, coronary artery disease (CAD), chronic obstructive pulmonary disease (COPD) and malignant tumor were all lower than 5.0%. The three most frequent symptoms of COVID-19 cases analyzed in this study were fever (82.9%), myalgia or fatigue (80.5%) and cough (63.4%). While, the three most frequent initial symptoms were myalgia or fatigue (80.5%), fever (73.2%), and cough (41.5%). Furthermore, other symptoms during course of disease included sputum production (39.0%), wheeze (34.1%), dyspnea (29.3%), chilly or shivering (29.3%), diarrhea (19.5%), dizziness (12.2%), somnipathy (12.2%), rhinobyon or running nose (9.8%), headache (4.9%), bloody sputum (2.4%), conjunctivitis (2.4%). (Table 1)

The mean white blood cell (WBC) count was 4.89 (± 1.83) ×10^9^/L. Cases with WBC count below 4.0×10^9^/L accounted for 36.6%. The mean lymphocyte count was 1.22 (± 0.58) ×10^9^/L. Cases with lymphocyte count below 1.0×10^9^/L accounted for 34.1%. Cases with procalcitonin below 0.2 ng/mL accounted for 97.6%. (Table 2)

**Table 2.**
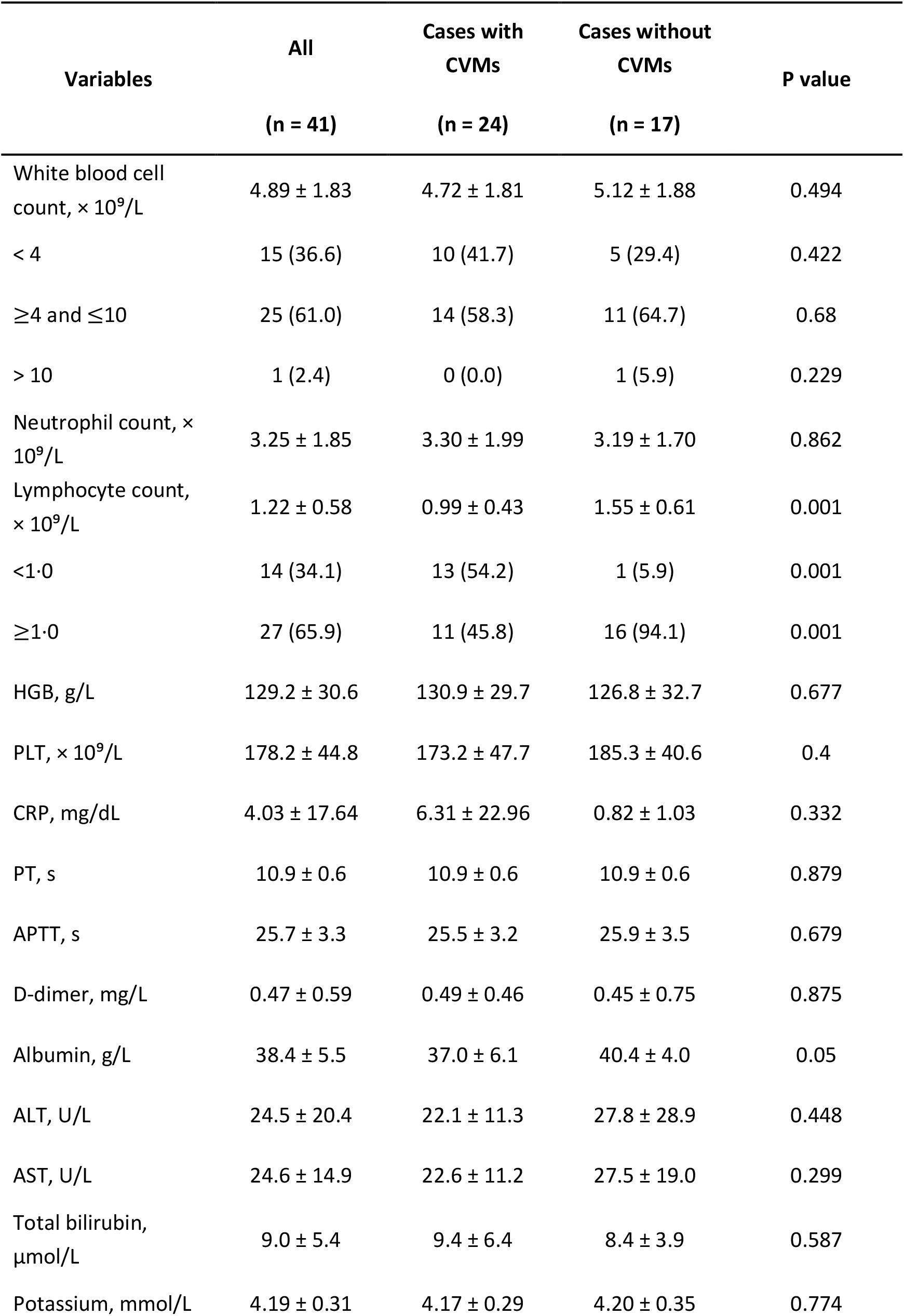
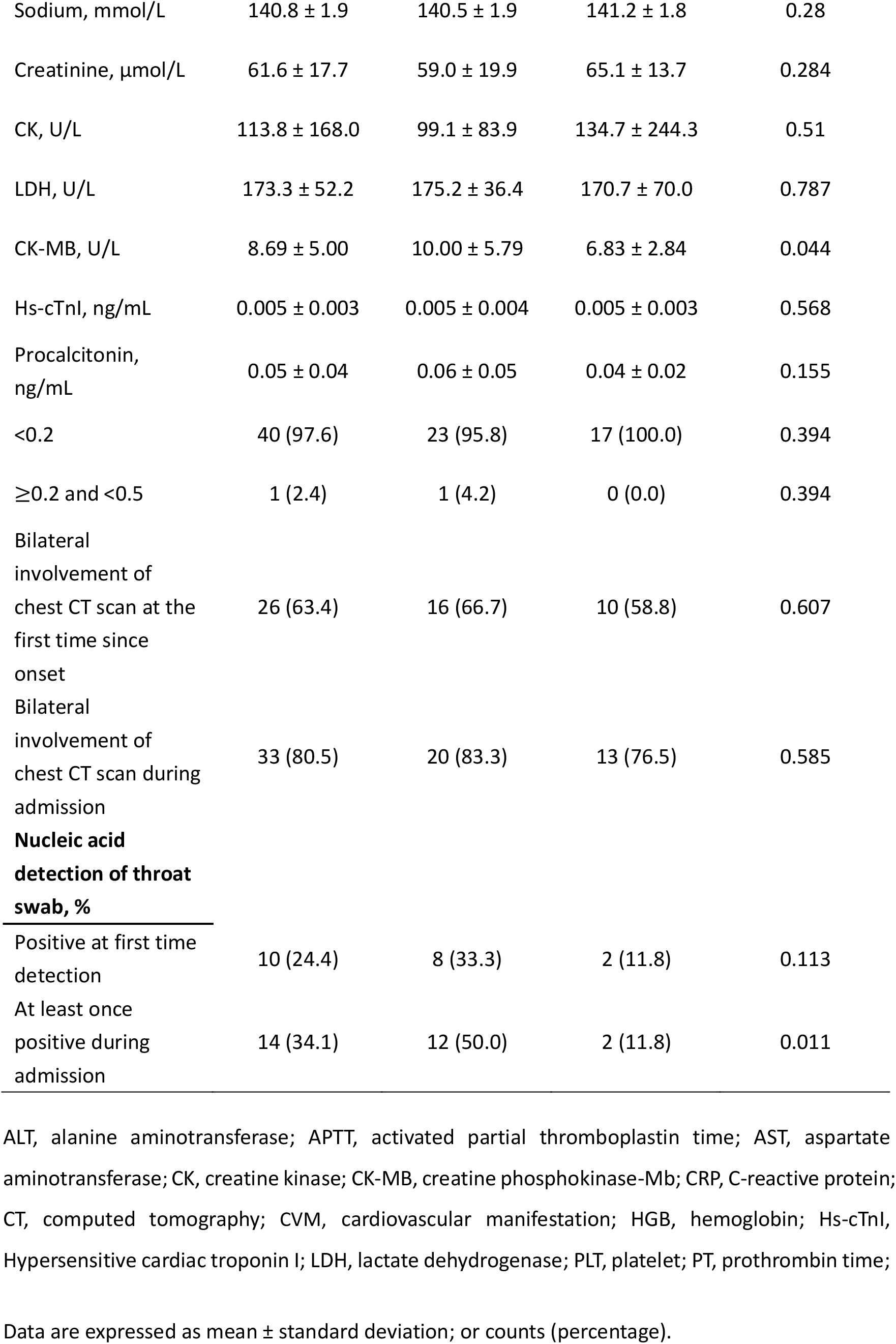
Baseline laboratory data and radiographic findings

| Variables | All<br>(n = 41) | Cases with<br>CVMs<br>(n = 24) | Cases without<br>CVMs<br>(n = 17) | P value |
| --- | --- | --- | --- | --- |
| White blood cell count, $\times 10^9/L$ | $4.89 \pm 1.83$ | $4.72 \pm 1.81$ | $5.12 \pm 1.88$ | 0.494 |
| < 4 | 15 (36.6) | 10 (41.7) | 5 (29.4) | 0.422 |
| $\geq 4$ and $\leq 10$ | 25 (61.0) | 14 (58.3) | 11 (64.7) | 0.68 |
| > 10 | 1 (2.4) | 0 (0.0) | 1 (5.9) | 0.229 |
| Neutrophil count, $\times 10^9/L$ | $3.25 \pm 1.85$ | $3.30 \pm 1.99$ | $3.19 \pm 1.70$ | 0.862 |
| Lymphocyte count, $\times 10^9/L$ | $1.22 \pm 0.58$ | $0.99 \pm 0.43$ | $1.55 \pm 0.61$ | 0.001 |
| <1.0 | 14 (34.1) | 13 (54.2) | 1 (5.9) | 0.001 |
| $\geq 1.0$ | 27 (65.9) | 11 (45.8) | 16 (94.1) | 0.001 |
| HGB, g/L | $129.2 \pm 30.6$ | $130.9 \pm 29.7$ | $126.8 \pm 32.7$ | 0.677 |
| PLT, $\times 10^9/L$ | $178.2 \pm 44.8$ | $173.2 \pm 47.7$ | $185.3 \pm 40.6$ | 0.4 |
| CRP, mg/dL | $4.03 \pm 17.64$ | $6.31 \pm 22.96$ | $0.82 \pm 1.03$ | 0.332 |
| PT, s | $10.9 \pm 0.6$ | $10.9 \pm 0.6$ | $10.9 \pm 0.6$ | 0.879 |
| APTT, s | $25.7 \pm 3.3$ | $25.5 \pm 3.2$ | $25.9 \pm 3.5$ | 0.679 |
| D-dimer, mg/L | $0.47 \pm 0.59$ | $0.49 \pm 0.46$ | $0.45 \pm 0.75$ | 0.875 |
| Albumin, g/L | $38.4 \pm 5.5$ | $37.0 \pm 6.1$ | $40.4 \pm 4.0$ | 0.05 |
| ALT, U/L | $24.5 \pm 20.4$ | $22.1 \pm 11.3$ | $27.8 \pm 28.9$ | 0.448 |
| AST, U/L | $24.6 \pm 14.9$ | $22.6 \pm 11.2$ | $27.5 \pm 19.0$ | 0.299 |
| Total bilirubin, $\mu\text{mol/L}$ | $9.0 \pm 5.4$ | $9.4 \pm 6.4$ | $8.4 \pm 3.9$ | 0.587 |
| Potassium, mmol/L | $4.19 \pm 0.31$ | $4.17 \pm 0.29$ | $4.20 \pm 0.35$ | 0.774 |
| Sodium, mmol/L | 140.8 ± 1.9 | 140.5 ± 1.9 | 141.2 ± 1.8 | 0.28 |
| Creatinine, µmol/L | 61.6 ± 17.7 | 59.0 ± 19.9 | 65.1 ± 13.7 | 0.284 |
| CK, U/L | 113.8 ± 168.0 | 99.1 ± 83.9 | 134.7 ± 244.3 | 0.51 |
| LDH, U/L | 173.3 ± 52.2 | 175.2 ± 36.4 | 170.7 ± 70.0 | 0.787 |
| CK-MB, U/L | 8.69 ± 5.00 | 10.00 ± 5.79 | 6.83 ± 2.84 | 0.044 |
| Hs-cTnI, ng/mL | 0.005 ± 0.003 | 0.005 ± 0.004 | 0.005 ± 0.003 | 0.568 |
| Procalcitonin, ng/mL | 0.05 ± 0.04 | 0.06 ± 0.05 | 0.04 ± 0.02 | 0.155 |
| <0.2 | 40 (97.6) | 23 (95.8) | 17 (100.0) | 0.394 |
| ≥0.2 and <0.5 | 1 (2.4) | 1 (4.2) | 0 (0.0) | 0.394 |
| Bilateral involvement of chest CT scan at the first time since onset | 26 (63.4) | 16 (66.7) | 10 (58.8) | 0.607 |
| Bilateral involvement of chest CT scan during admission | 33 (80.5) | 20 (83.3) | 13 (76.5) | 0.585 |
| **Nucleic acid detection of throat swab, %** |  |  |  |  |
| Positive at first time detection | 10 (24.4) | 8 (33.3) | 2 (11.8) | 0.113 |
| At least once positive during admission | 14 (34.1) | 12 (50.0) | 2 (11.8) | 0.011 |

ALT, alanine aminotransferase; APTT, activated partial thromboplastin time; AST, aspartate aminotransferase; CK, creatine kinase; CK-MB, creatine phosphokinase-Mb; CRP, C-reactive protein; CT, computed tomography; CVM, cardiovascular manifestation; HGB, hemoglobin; Hs-cTnI, Hypersensitive cardiac troponin I; LDH, lactate dehydrogenase; PLT, platelet; PT, prothrombin time;
Data are expressed as mean ± standard deviation; or counts (percentage).

There were 26 of the 41 enrolled patients (63.4%) who showed bilateral involvement of chest computed tomography (CT) scan at the first time since onset. And cases with bilateral involvement of chest CT scan during admission accounted for 80.5%, lower than the percentage previously reported [11, 12] (Table 2, Figure 1).

**Figure 1.**
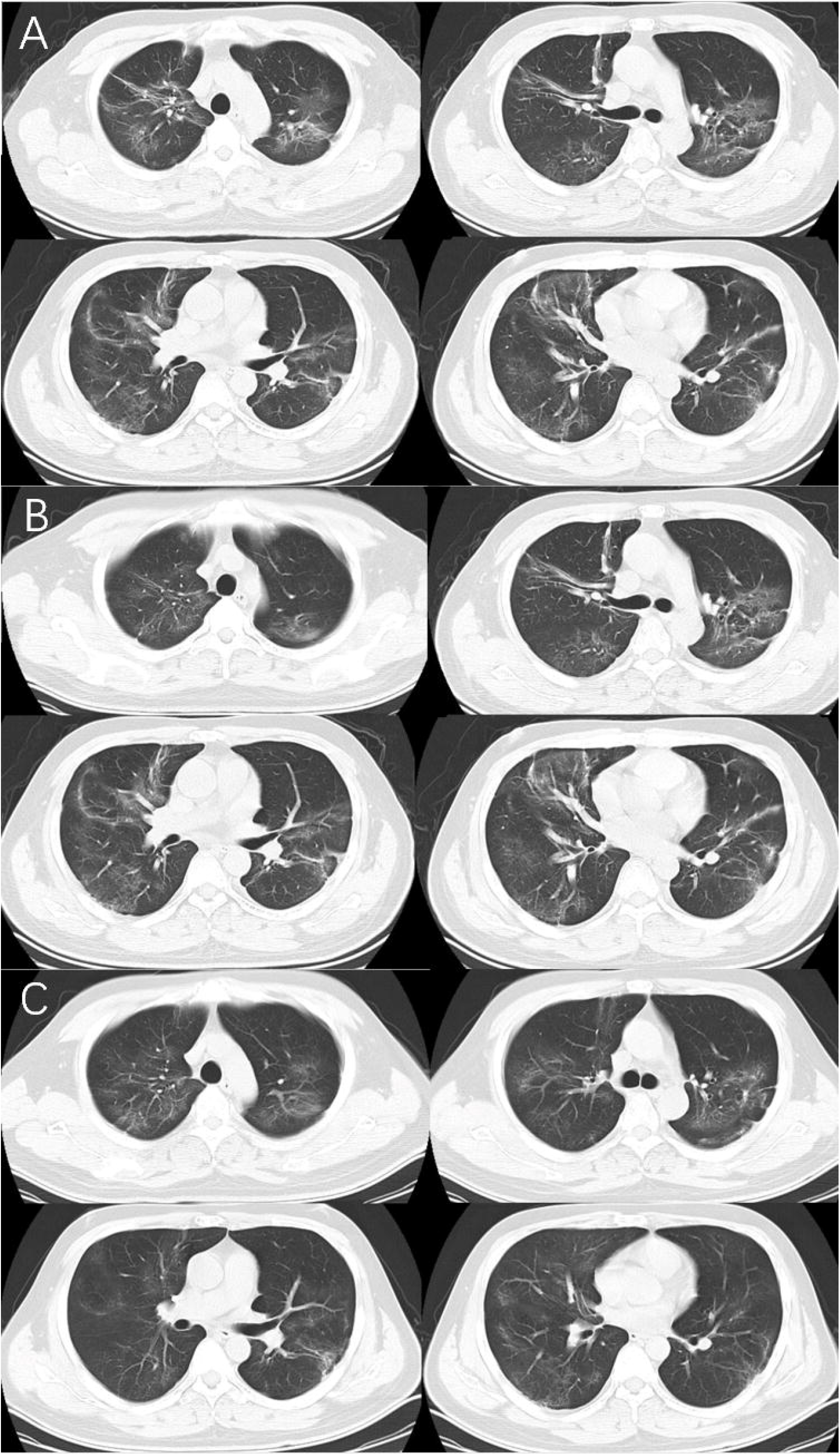
Chest computed tomographic images of a 32-year-old male doctor on 120 Ambulance infected with SARS-CoV-2 A, Chest computed tomographic images obtained on February 1, 2020, show ground glass opacity in both lungs on day 19 after symptom onset. B, Images taken on February 9, 2020, show the slowly absorption of bilateral ground glass opacity after about 4 weeks treatment. C, Images taken on February 16, 2020, show yet the poorly absorption of bilateral ground glass opacity after about 5 weeks treatment. The patients received mesenchymal stem cell therapy on February 17, 2020.

All 41 cases received at least twice nucleic acid detection of throat swab during admission. Only those with normal temperature for more than 3 days, improved respiratory symptoms, obvious absorption of pulmonary inflammation on radiography, as well as twice negative nucleic acid detection of throat swab with interval more than 24 hours, can be released from quarantine and discharged, according to the Diagnosis and Treatment Scheme of COVID-19 (the fifth trial edition) by National Health Commission and National Administration of Traditional Chinese Medicine of China [15]. Cases with positive detection at first time accounted for 24.4%, while those with at least once positive result during admission accounted for 34.1%. (Table 2)

### Treatment Situation and In-hospital Outcomes

There were 40 cases (97.6%) received at least one kind of antiviral drugs empirically including oseltamivir, ribavirin, arbidol or lopinavir/ritonavir (Kaletra), which were accessible clinically in China. Antibiotics or anti-fungus drugs were applied in 95.1% cases regarding to confirmed secondary infection or preventing secondary infection in relatively severe cases. Corticosteroid was used in 78.0% cases to control immune overreaction. Human albumin was applied when serum albumin decreased below 30g/L. Immunoglobulin was also applied empirically, as well as thymosin. Cases who received Chinese traditional medicine accounted for 19.5%, including patent medicine and prescription. There were 4 cases as of March 3 received mesenchymal stem cell therapy after informed consent. One of them firstly consented this empirical therapy on day 35 after symptom onset as pulmonary inflammation poorly absorbed on CT scan (Figure 1). There were 23 cases (56.1%) underwent respiratory support, including oxygen therapy through only common nasal catheter, face mask, both common nasal catheter and face mask simultaneously, high flow nasal cannula and mechanic ventilation. Only one case suffered from irreformable hypoxemia, received noninvasive ventilator support, and later transferred to ICU for tracheal intubation. (Table 3)

**Table 3.** Treatment situation

| Variables | All<br>(n = 41) | Cases with CVMs<br>(n = 24) | Cases without<br>CVMs<br>(n = 17) | P value |
| --- | --- | --- | --- | --- |
| Antiviral drugs, % | 40 (97.6) | 24 (100.0) | 16 (94.1) | 0.229 |
| Kaletra, % | 16 (39.0) | 11 (45.8) | 5 (29.4) | 0.288 |
| Antibiotics, % | 39 (95.1) | 23 (95.8) | 16 (94.1) | 0.802 |
| Corticosteroid, % | 32 (78.0) | 20 (83.3) | 12 (70.6) | 0.331 |
| Human albumin or<br>immunoglobulin, % | 33 (80.5) | 20 (83.3) | 13 (76.5) | 0.585 |
| Thymosin, % | 16 (39.0) | 10 (41.7) | 6 (35.3) | 0.68 |
| Chinese traditional<br>medicine, % | 8 (19.5) | 6 (25.0) | 2 (11.8) | 0.292 |
| Mesenchymal<br>stem cell, % | 4 (9.8) | 4 (16.7) | 0 (0.0) | 0.076 |
| **Respiratory**<br>**support, %** |  |  |  |  |
| Oxygen therapy | 23 (56.1) | 19 (79.2) | 4 (23.5) | <0.001 |
| Common nasal<br>catheter | 23 (56.1) | 19 (79.2) | 4 (23.5) | <0.001 |
| Face mask | 7 (17.1) | 6 (25.0) | 1 (5.9) | 0.109 |
| High flow nasal<br>cannula | 8 (19.5) | 7 (29.2) | 1 (5.9) | 0.064 |
| Noninvasive<br>ventilator | 1 (2.4) | 1 (4.2) | 0 (0.0) | 0.394 |
| Invasive<br>mechanical<br>ventilation | 1 (2.4) | 1 (4.2) | 0 (0.0) | 0.394 |
| ECMO | 1 (2.4) | 1 (4.2) | 0 (0.0) | 0.394 |

CVM, cardiovascular manifestation; ECMO, extracorporeal membrane oxygenation
Data are expressed as counts (percentage).

There was none in-hospital death, and none suffered from acute cardiac injury or AKI during admission as to March 3,2020. Two cases were diagnosed as RF or ARDS (4.9%). Most cases featured non-severe course of disease, accounted for 95.1% according to official criteria [15, 25]. One case was recognized as acute myocyte injury (2.4%). Cases suffered from acute hepatic injury accounted for 19.5%, who all received conventional liver protecting treatment including Essentiale or diammonium glycyrrhizinate. Secondary infection happened in 20 cases (48.8%). Only one case (2.4%) transferred to the negative pressure respiratory ward of ICU in Wuhan Chest Hospital for invasive mechanical ventilation and received extracorporeal membrane oxygenation (ECMO) support on the next day due to rapid deterioration. The total in-hospital adverse event rate was 53.7%. There are still 5 cases not up to the discharge criteria on the final date of follow-up, despite of clinical improvement and nucleic acid detection of throat swab turning the negative [15, 25]. (Table 4)

**Table 4.** In-hospital outcomes

|  | All<br>(n = 41) | Cases with CVMs<br>(n = 24) | Cases without<br>CVMs<br>(n = 17) | P value |
| --- | --- | --- | --- | --- |
| In-hospital adverse events | 22 (53.7) | 18 (75.0) | 4 (23.5) | 0.001 |
| RF or ARDS | 2 (4.9) | 2 (8.3) | 0 (0.0) | 0.222 |
| Acute myocyte injury | 1 (2.4) | 0 (0.0) | 1 (5.9) | 0.229 |
| Acute hepatic injury | 8 (19.5) | 7 (29.2) | 1 (5.9) | 0.064 |
| Secondary infection | 20 (48.8) | 17 (70.8) | 3 (17.6) | 0.001 |
| Transfer to ICU | 1 (2.4) | 1 (4.2) | 0 (0.0) | 0.394 |
| Invasive mechanical ventilation | 1 (2.4) | 1 (4.2) | 0 (0.0) | 0.394 |
| ECMO | 1 (2.4) | 1 (4.2) | 0 (0.0) | 0.394 |

ARDS, acute respiratory distress syndrome; CVM, cardiovascular manifestation; ECMO, extracorporeal membrane oxygenation; RF, respiratory failure; ICU, intensive care unit; Data are expressed as counts (percentage).

### A Comparative Analysis

Among 41 cases analyzed, patients with CVMs and those without CVMs accounted for 58.5% and 41.5%, respectively. Compared with cases without CVMs, patients with CVMs were presented with lower baseline lymphocyte count (0.99 ± 0.43 and 1.55 ± 0.61, *P*<0.001), more who had at least once positive nucleic acid detection of throat swab during admission (50.0% and 11.8%, *P*=0.011), and more received oxygen support (79.2% and 23.5%, *P*<0.001). The in-hospital adverse events happened more in patients with CVMs group (75.0% and 23.5%, *P*=0.001). Multivariable logistic regression model indicated that, after adjustment of the possible baseline confounders, concomitant with CVMs was not independently associated with in-hospital adverse events (OR 2.23, 95%CI 0.24-20.27, *P* = 0.478) in COVID-19 patients. Baseline lymphocyte count showed a tendency of significance between the two groups in model (OR 0.15, 95%CI 0.02-1.26, *P* = 0.081). (Table 5)

**Table 5.** Multivariate logistic regression analysis

| Covariates | In-hospital adverse events |  |
| --- | --- | --- |
|  | OR (95%CI) | P value |
| Cases with CVMs | 2.23 (0.24, 20.27) | 0.478 |
| Sputum production | 0.48 (0.06, 3.76) | 0.485 |
| Somnipathy | 0.52 (0.03, 8.28) | 0.645 |
| Lymphocyte count | 0.15 (0.02, 1.26) | 0.081 |
| Albumin | 0.89 (0.70, 1.15) | 0.377 |
| CK-MB | 1.10 (0.80, 1.50) | 0.565 |
| At least once positive during admission | 3.19 (0.35, 29.24) | 0.305 |
| Oxygen therapy | 4.02 (0.53, 30.31) | 0.177 |

CVM, cardiovascular manifestation; CK-MB, creatine phosphokinase-Mb
Data are expressed as counts (percentage).
Covariates were those possible confounders, showing a significant difference or a tendency of significant difference between 2 groups in baseline analysis.

## Discussion

The analyzed population is characterized as younger age, less comorbidities than common community patients [11, 12], and mostly featured non-severe course of disease, which is probably an important clinical characteristic of COVID-19 infection of hospital staff. As of March 3, 2020, only one case underwent invasive mechanic ventilation and ECMO support. Other cases all presented markedly improvement of symptoms and findings on chest CT images, due to aggressive and comprehensive medical treatment, although they also suffered from many adverse events during hospitalization, like secondary infection or acute hepatic injury. This result is consistent with the situation of infected hospital workers in a large data reported by the NCP Emergency Response Epidemiology Team of China [13].

Our data showed that, the infection of SARS-CoV-2 in hospital staff mostly had definite or suspected history of patient contact, which was consistent with most reports. The transmission of SARS-CoV-2 mainly characterized as confined space and close contact. No other countries around the world experiences a similar time in great favor of transmission of SARS-CoV-2, like the Spring Festival and the Spring Festival travel season in China. During this period of year in China, people’ s lives are full of behavior styles characterized as confined space and close contact, including air or railway transportation for return home or travel, various familial or public get-togethers. Nosocomial infection is another key feature of the outbreak. At the early time of prevalence, a large number of suspected patients squeezed into fever clinics or emergency departments of hospitals in Wuhan, in which health workers had not well prepared in terms of both protective supplies and awareness. Meanwhile, patients in incubation period of COVID-19 underwent elective surgeries who were diagnosed after procedures made nosocomial infection quite a few on the early stage of the outbreak. As continually support of protective materials from other areas into Hubei Province, as well as improved knowledge of protection, data with decreased risk of nosocomial infection are expected.

Xu et al reported the pathological findings of a 50-year-old male with COVID-19, died of ARDS and sudden cardiac arrest. They found that there were a few interstitial mononuclear inflammatory infiltrates, but no other substantial damage in the heart tissue [20]. The result implied the possibility of coexisting myocarditis, as part of systemic inflammatory response syndrome. And the persistent hypoxemia may directly cause myocardial suppression, possibly related to sudden cardiac arrest [21]. In other words, the CVMs of COVID-19 maybe more associated with the systemic immune reaction, rather than direct damage of heart by virus attack. And these mechanisms maybe explain the concomitant CVMs of COVID-19 were not independently associated with adverse in-hospital outcomes to some extent.

In this study, there were about 20.5% of cases with non-severe course of COVID-19 suffered from acute hepatic injury, needed liver protection treatment. And it probably should not be totally attributed to drug-induced liver injury and aroused more concern on COVID-19 itself. Xu et al reported the liver biopsy specimens of the patient with COVID-19 showed moderate microvascular steatosis and mild lobular and portal activity, indicating the injury could have been caused by either SARS-CoV-2 infection or drug-induced liver injury [20]. Pathological findings were also in favor of more attention to the hepatic injury in the pathogenesis of COVID-19.

There is no specific antiviral drug for SARS-CoV-2 infection up to now. Expectation mainly comes from evidences that remdesivir and chloroquine are highly effective in the control of 2019-nCoV infection in vitro, while their cytotoxicity remains in control [22]. It was reported that remdesivir, a nucleotide analogue prodrug, in previously clinical development for treatment of Ebola virus disease, exhibited broad-spectrum anti-coronavirus (CoV) activity. It could inhibit severe acute respiratory syndrome coronavirus (SARS-CoV) and Middle East respiratory syndrome coronavirus (MERS-CoV) replication in multiple in vitro systems, and was also effective against bat CoVs, prepandemic bat CoVs, and circulating contemporary human CoV in primary human lung cells. In a mouse model of SARS-CoV pathogenesis, prophylactic and early therapeutic administration of remdesivir significantly reduced lung viral load and improved clinical signs of disease as well as respiratory function [23]. These preclinical evidences, together with successful clinical cases treated by remdesivir in USA and France [24] made us looking forward to the final result of the randomized controlled trial unblinding before long in Wuhan. Meanwhile, chloroquine has been recently written into official recommendation for empirical therapy of COVID-19 for its adequate safety data in human [25]. It is a cheap and safe drug that has been used as an antimalarial for more than 70 years. Combination of lopinavir and ritonavir (Kaletra) among SARS-CoV patients was reported a substantial clinical benefit [26]. As it is available in the designated hospital as anti-human immunodeficiency virus drug, a randomized controlled trial has been initiated quickly to assess the efficacy and safety of it [11]. Other medication as newly recommended officially are mostly empirical. Experience came from the outbreak of SARS-CoV 17 years ago. From empirical medicine to evidence-based medicine, it is the only way which must be passed that forms comprehensive knowledge about the new virus and the new disease. And for this, the Chinese people have paid a heavy price, deserved to be remembered.

### Limitation

Firstly, the cases enrolled in this study were only hospital staff. The conclusion cannot be extrapolated to common community patients. Secondly, as the suddenness of the outbreak, the vast patient volume in hospitals and shortage of healthcare personnel, it is hard to obtain large clinical data. The sample is not large enough for observation of mortality in severe cases of COVID-19. The multivariate model analysis had limitation due to the sample size. And we cannot figure up how the research variable effected each adverse in-hospital event. Lastly, the in-hospital outcomes were monitored up to March 3, 2020, the final date of follow-up. The incomplete follow-up data will be made up constantly. Besides, we are trying to collect the data of the severe case who transferred to other hospital. Nevertheless, this is first-hand data in terms of hospital staff infection in Wuhan, and the first analysis comparing the in-hospital outcomes of COVID-19 between cases with CVMs and those without CVMs, in anticipation of finding the risk factors in favor of severe conversion of non-severe case.

## Conclusions

Most hospital staff with COVID-19 had history of patient contact, featured a non-severe course of disease. COVID-19 patients with CVMs suffered from more in-hospital adverse events than those without CVMs. But the concomitant CVMs were not independently associated with in-hospital adverse events in patients with COVID-19.

## Data Availability

All data, models, or code generated or used during the study are available from the corresponding author by request.

## Acknowledgements

We thank all the patients; the nurses and clinical staff who are providing care for the patients. We thank professors in Beijing Fuwai Hospital, Chinese Academy of Medical Sciences and Peking Union Medical College, for data review.

## Authors’ contributions

RL contributed to all aspects of this study, including study concept and design, data review and arrangement, statistical analysis and interpretation, drafting and revising the report, and funding. HZ, XYM and JMZ contributed to study concept, data acquisition and verification, and funding. HP and NX contributed to data acquisition. OX and JLZ contributed to professional consultation. All authors have approved the final article.

